# Comparison of in-hospital mortality and clinical outcomes among patients greater than and less than 80 years undergoing TAVR

**DOI:** 10.1101/2022.01.11.22269040

**Authors:** Mukunthan Murthi, Sujitha Velagapudi, Bharosa Sharma, Emmanuel Akuna, Olisa Ezegwu Kingsley, Dae Yong Park, Ramtej Atluri, Ufuk Vardar

## Abstract

**Background:** Transcatheter aortic valve replacement (TAVR) procedure has been increasingly utilized in the management of aortic stenosis in the elderly. We sought to assess the hospital outcomes and major adverse events of TAVR in patients ≥80 years old compared to those <80 years.

**Methods:** We performed a retrospective observational study using the National Inpatient Sample - 2018. We divided TAVR patients into two cohorts based on age being ≥80 years and <80 years old. The primary outcomes were the comparison of in-hospital mortality and major adverse events (MAE) in the cohorts.

**Results:** We identified 63,630 patients who underwent TAVR procedures from January 1 to December 31, 2018. Among them, 35, 115(55%) were ≥80 years and 28,515(45%) were <80 years of age. There was a higher rate of post-procedure in-hospital mortality rate in patients ≥80 years (1.6% vs. 1.1%, aOR=1.56, [CI 1.13 -2.16], p=0.006). They also had higher rates of pacemaker insertion compared to those <80 years old (7.4 vs 6.5%, aOR=1.17 [CI 1-1.35], p=0.03). On subgroup analysis, the rates of MAE were not different between the two cohorts (23.8 vs 23.4, p=0.09) but patients ≥80 years who experienced MAE had higher in-hospital mortality (5.7 vs 4.3%, aOR=1.58 [CI=1.08-2.32], p=0.01) and shorter length of hospital stay (7.2 vs 8.7 days, p=0.03) compared to those <80. Anemia, liver disease, chronic kidney disease and previous stroke were associated with higher odds of in-hospital MAE in both groups.

**Conclusion:** This study shows that in patients undergoing TAVR, those ≥80 years old had higher in-hospital mortality and higher rates of pacemaker insertion compared to those < 80 years. The rates of MAE were not significantly different between the two groups.

## 1. Introduction

Aortic stenosis is the leading cause of left ventricular outflow tract obstruction, notably in the elderly. The most common causes of aortic stenosis include bicuspid valve, calcification, and rheumatic heart disease(1). About 5% of the population over 65 years of age have aortic stenosis and the percentage increases exponentially with older age. In addition, the prevalence of severe aortic stenosis also increases with age, with 3.4 to 4.3% of adults over 75 years having severe aortic stenosis (2, 3).

For many years, surgical aortic valve replacement has been the standard of care for severe symptomatic aortic stenosis until the advent of transcatheter aortic valve replacement (TAVR). TAVR has been the alternative treatment option in patients considered unsuitable for surgery (4). From the initial approval for patients with severe AS and prohibitive operative risk, it is currently utilized even for severe aortic stenosis and low-risk patients(5, 6).

Several studies have compared the outcomes of TAVR compared to Surgical aortic valve replacement (SAVR). Studies have also compared these outcomes specifically in octogenarians(7, 8) and Nonagenarians(9). The results of these studies favor the use of TAVR in patients older than 80 years and this is reflected in the recent American heart associated/American College of Cardiology guidelines (10). However, there are limited data comparing the difference in outcomes of TAVR between patients more than 80 years and less than 80 years old using large population databases. Therefore, we wanted to analyze the demographic characteristics and in-hospital outcomes after the TAVR procedure in patients ≥80 years old compared to those <80 years. We also wanted to investigate the factors independently associated with major adverse events (MAE) in these groups.

## 2. Materials and Methods

### 2.1. Design and Data source

We performed a retrospective study involving adult hospitalizations undergoing the TAVR procedure in the USA in 2018. We extracted data from the National Inpatient Sample for the year 2018. The NIS is the largest publicly available all-payer inpatient admission database in the USA. It is developed by the Agency for Healthcare Research and Quality (AHRQ) Healthcare Cost and Utilization Project (HCUP) State Inpatient Databases (SID). This dataset contains discharge information for over 7 million discharges annually with data stratified as a weighted sample. Discharge weights were calculated using post-stratification on hospital characteristics (census region, urban-rural location, teaching status, bed size, and hospital control) and patient characteristics (sex and five age groups [0, 1-17, 18-44, 45-64, and 65 and older]). Since the NIS does not include individual patient identifiers it does not require approval from the Cook County Health Institutional Review Board. This manuscript conforms with the STROBE statement for observational studies.

### 2.2. Study population and variables

We identified patients who underwent TAVR procedure in the year 2018 using the International Classification of Diseases, Tenth Revision, Clinical Modification (ICD□10□CM) procedure codes (Table S1 and Table S2). We further divided these patients into two groups based on age being ≥80 years and <80 years old. The NIS dataset includes variables on patient demographics, including age, gender, race, median household income, and type of insurance. It also contains hospital-level data including hospital bed size, teaching status, and location. Comorbidities were identified using ICD-10 codes as well as Sundararajan’s adaptation of the modified Deyo’s Charlson comorbidity index (CCI)(Table S3) (11).

### 2.3. Measures of Outcome

The primary outcomes were the comparison of mortality and in-hospital major adverse events (MAE) in patients with TAVR procedure stratified based on age. We included post-procedural hemorrhage, cardiac complications, acute kidney injury (AKI), stroke, and TIA as MAE. Secondary outcomes included rates of pacemaker insertion, assessment of the mean length of hospital stay (LOS), total hospital charges (THC), and independent predictors of MAE.

### 2.4. Statistical analysis

We analyzed the data using Stata® Version 16 software (StataCorp, Texas, USA). We conducted all the analyses using the weighted samples for national estimates in accordance with HCUP guidelines. We calculated comorbidities as proportions of the cohorts and used the Chi-square test for comparison. We used univariate regression to identify variables affecting primary outcomes. We included those variables having a p-value < 0.1 in the final multivariate regression model. Variables identified to be significant by literature review were also forced into the model. Subsequently, we ran a multivariate cox regression analysis to identify independent predictors of MAE with p-values <0.05 set as the threshold for statistical significance. For MAE analysis in all TAVR patients, these variables included age ≥80 years, Gender, Race, Zip-code wise median household income, region of hospital, insurance status, Charlson comorbidity category, COPD, previous stroke, Obesity, Diabetes mellitus, peripheral vascular disease, Liver disease, smoking history, and anemia. For MAE analysis in patients ≥80, the variables used in the multivariate analysis include age, Gender, Race, Charlson comorbidity category, COPD, previous stroke, hypertension, diabetes mellitus, heart failure, chronic kidney disease, Liver disease, hemodialysis, smoking history, and anemia. For MAE analysis for patients <80, variables for multivariate analysis included age, Gender, Race, Charlson comorbidity category, insurance status, COPD, previous stroke, hypertension, diabetes mellitus, heart failure, chronic kidney disease, Liver disease, smoking history, and anemia.

## 3. Results

We identified 63,630 patients who underwent the TAVR procedure from January 1 to December 31, 2018. Among them, 35, 115(55%) were ≥80 years and 28,515(45%) were <80 years of age. Among patients ≥80 and <80 years of age, the mean age was 85(SE ±0.04) vs 71 years (SE±0.11) (p<0.001) and the proportion of females were 48% vs 44% respectively (p=0.0002). European whites formed the major proportion of both groups. The number of patients with ≥3 comorbidities was higher in those <80 years old (51.2 vs 59.6%, p<0.001). A larger proportion of patients <80 years old were privately insured compared to those ≥80 years old (3.8 vs 12%, p>0.001). All other baseline characteristics are shown in Table 1 and Figure 1.

**Table 1:** Clinical and demographic characteristics of TAVR patients.

| **Patient characteristics** | **≥80** | **<80** | **P value** |
| --- | --- | --- | --- |
| **Subjects (n,%)** | 35,115(55%) | 28,515(44%) |  |
| **Mean age (in years) (SE)** | 85±0.04 | 71±0.11 | <0.001 |
| **Female (%)** | 48 | 44 | 0.0002 |
| **Race (%)** |  |  | <0.001 |
| **White** | 88 | 83 |  |
| **Black** | 3 | 6.2 |  |
| **Hispanic** | 5 | 6.7 |  |
| **Other** | 4 | 4 |  |
| **Charlson Category**<br>**(No. of comorbidities) (%)** |  |  | <0.001 |
| **0** | 7.4 | 5 |  |
| **1** | 20.6 | 15.6 |  |
| **2** | 20.6 | 19.6 |  |
| **≥3** | 51.2 | 59.6 |  |
| **Zip-code wise median income (in \$) (%)** | | | <0.001 |
| **1 - 45,999** | 18.3 | 24.7 |  |
| **46,000 - 58,999** | 25.2 | 25.9 |  |
| **59,000 - 78,999** | 27.6 | 26.7 |  |
| **79,000+** | 28.8 | 23.1 |  |
| **Insurance (%)** |  |  | <0.001 |
| **Medicare** | 95.5 | 84 |  |
| **Medicaid** | 0.3 | 3 |  |
| **Private** | 3.8 | 12 |  |
| **Other** | 0.1 | 0.7 |  |
| ***Hospital region (%)*** |  |  | <0.001 |
| **Northeast** | 23.2 | 19.1 |  |
| **Midwest** | 22 | 22.4 |  |
| **South** | 32.6 | 38 |  |
| **West** | 21.9 | 20.3 |  |
| ***Hospital Bed-size (%)*** |  |  | 0.17 |
| **Small** | 6.9 | 7.6 |  |
| **Medium** | 21 | 19.5 |  |
| **Large** | 72 | 72.8 |  |
| ***Location/teaching status of hospital (%)*** |  |  | 0.51 |
| **Rural** | 0.8 | 0.6 |  |
| **Urban non-teaching** | 9.1 | 8.8 |  |
| **Urban teaching** | 90 | 90.5 |  |
| ***Comorbidities (%)*** |  |  |  |
| **COPD** | 19.4 | 28.2 | <0.001 |
| **Stroke** | 15 | 11.8 | <0.001 |
| **Hypertension** | 17.9 | 17.9 | 0.97 |
| **PVD** | 9.6 | 8.1 | 0.003 |
| **Diabetes** | 28.9 | 44.6 | <0.001 |
| **Obese** | 10.9 | 27.6 | <0.001 |
| **Heart failure** | 72.7 | 73.7 | 0.25 |
| **CKD** | 35.8 | 34.9 | 0.32 |
| **Liver disease** | 1.6 | 5.7 | <0.001 |
| **Hemodialysis** | 1.2 | 5 | <0.001 |
| **Smoking** | 34.8 | 36.8 | 0.02 |
| **Anemia** | 32.1 | 33.8 | 0.04 |

**Figure 1:**
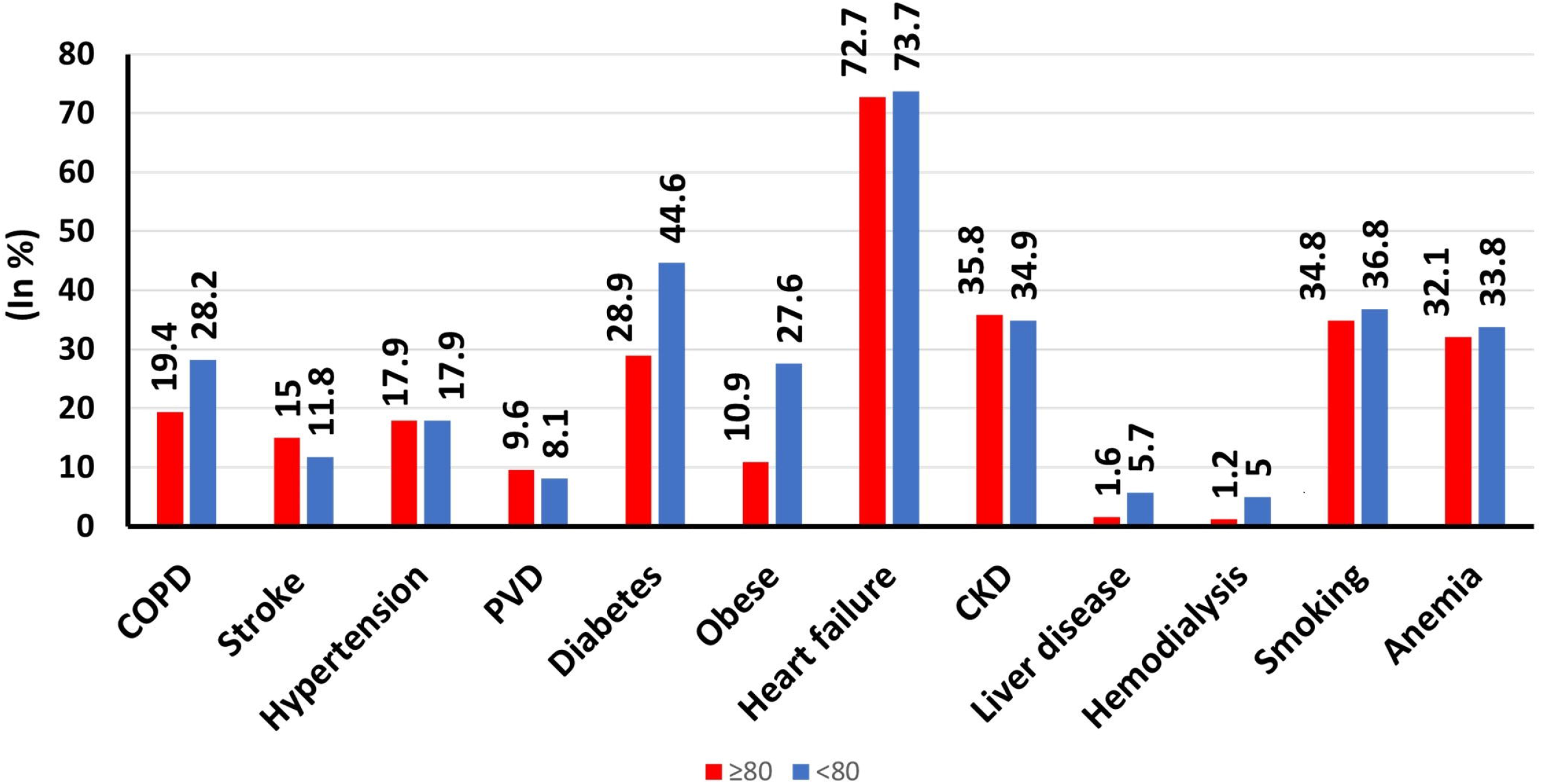
Graph showing the prevalence of comorbidities among TAVR patients ≥80 years vs < 80 years

### 3.1. Comparison of TAVR patients ≥80 years vs < 80 years

The in-hospital mortality rate for patients ≥80 years and < 80 years of age was 1.6% and 1.1% respectively (aOR=1.56, [CI 1.13 -2.16], p=0.006). The rates of MAE were not different in patients ≥80 years of age compared to those <80 years (23.8 vs 23.4, p=0.09). There were no significant differences in the length of stay (3.7 vs 4.2 days, p=0.26) and total hospital charges ($214,919 vs $220,681, p=0.42) between patients ≥80 years and < 80 years of age (Table 2). Patients ≥80 years had higher rates of pacemaker insertion (7.4 vs 6.5%, aOR=1.17 [CI 1-1.35], p=0.03). On multivariate regression analysis, age ≥80 years was not independently associated with increased MAE in TAVR patients. Figure 2 shows the independent factors associated with MAE.

**Table 2:** In hospital outcomes and Major adverse events in TAVR patients stratified by age

| **Variable (%)** | **≥80 years age** | **<80 years age** | **p- value<sup>#</sup>** |
| --- | --- | --- | --- |
| **MAE** | 23.8 | 23.4 | 0.09 |
| **Post procedural hemorrhage** | 1.7 | 1.7 | 0.54 |
| **Cardiac complications** | 13.2 | 12.9 | 0.51 |
| **AKI** | 10.3 | 11.3 | 0.14 |
| **Stroke & TIA** | 4.2 | 3.5 | 0.26 |
| **Pacemaker insertion** | 7.4 | 6.5 | 0.03 |
| **Died** | 570(1.6%) | 335(1.1%) | 0.006 |
| **Length of stay (in days)** | 3.7(CI 3.5-3.8) | 4.2 (CI 4-4.4) | 0.26 |
| **Total hospital discharges (in \$)** | 214,919 | 220,681 | 0.42 |

<sup>#</sup>Multivariate analysis
MAE= Major adverse events, TIA= Transient ischemic attack, AKI= Acute kidney injury

**Figure 2:**
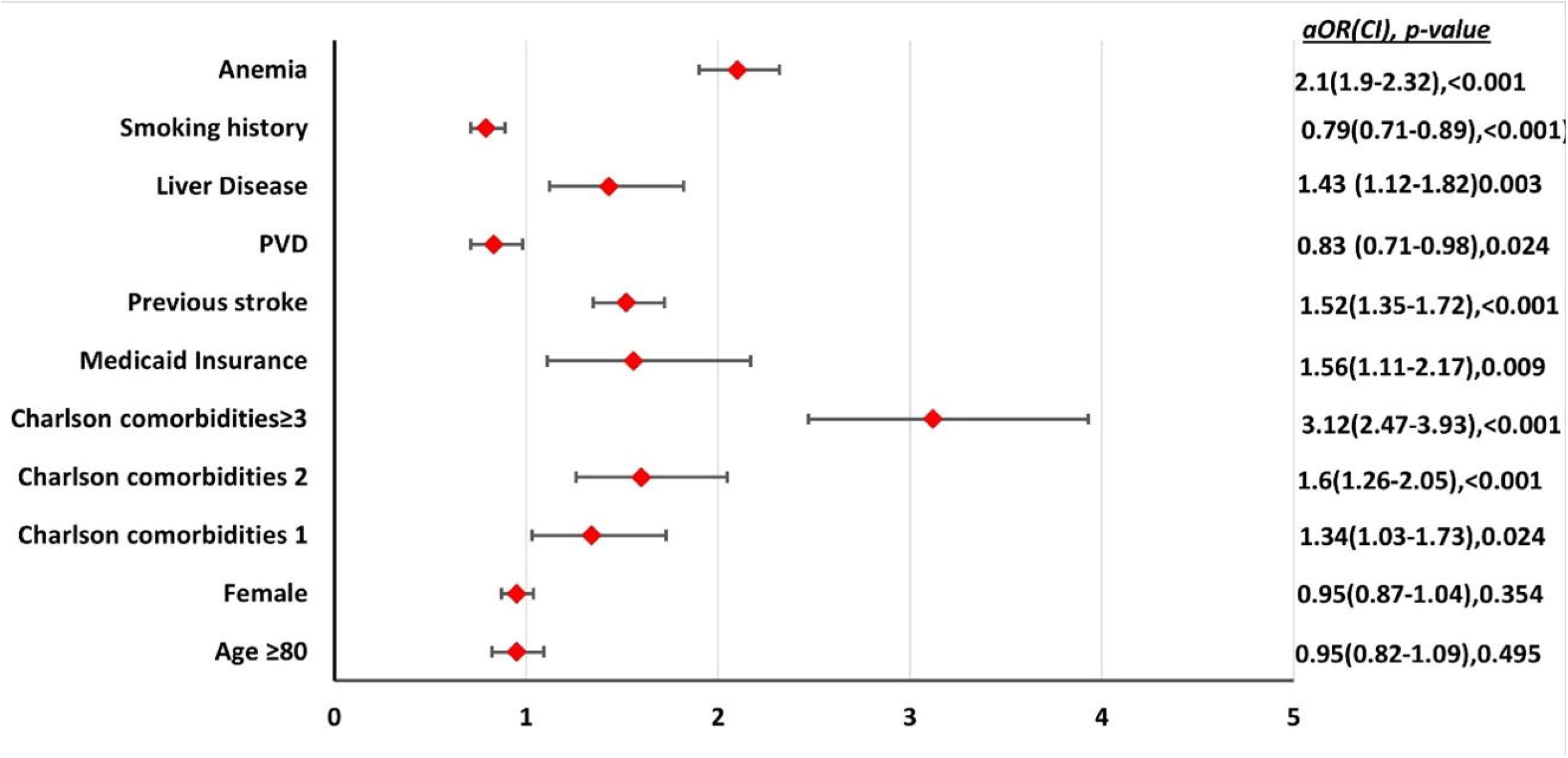
Independent factors associated with MAE in patients undergoing TAVR. PVD-Peripheral vascular Disease

### 3.2. Comparison of TAVR patients ≥80 years with and without MAE

The mean age for patients with and without MAE was 85.5(SE±0.08) vs 85.1(SE±0.04) respectively (<0.001). Patients with MAE had significantly higher comorbidities compared to those without MAE (Table 3). Anemia (aOR-2.12), Liver disease (aOR-1.57), CKD (aOR-1.34), history of stroke (aOR-1.54) and higher number of comorbidities (aOR-1.97) were independently associated with higher odds of MAE (Figure 3). Increasing age was also associated with worse outcomes (aOR-1.03).

**Table 3:**

| Patient characteristics | ≥80<br>MAE | ≥80<br>No MAE | P value | <80<br>MAE | <80<br>No MAE | P value | P value<br>* |
| --- | --- | --- | --- | --- | --- | --- | --- |
| Mean age (in years)<br>(SE) | 85.5±0.08 | 85.1±0.04 | <0.001 | 71.2±0.19 | 71.4±0.13 | <0.001 | <0.001 |
| Female (%) | 47.3 | 48.1 | 0.55 | 44.6 | 44.7 | 0.92 | 0.13 |
| Race (%) |  |  | 0.31 |  |  | 0.10 | 0.01 |
| White | 87.6 | 88.2 |  | 82.1 | 83.4 |  |  |
| Black | 3.3 | 3.1 |  | 6.2 | 6.3 |  |  |
| Hispanic | 6.1 | 4.8 |  | 1.9 | 6.1 |  |  |
| Other | 2.7 | 3.1 |  | 3.3 | 3.7 |  |  |
| Charlson Category<br>(No. of comorbidities)<br>(%) |  |  | <0.001 |  |  | <0.001 | <0.001 |
| 0 | 3.9 | 8.5 |  | 1.9 | 6 |  |  |
| 1 | 13.5 | 22.9 |  | 8.6 | 17.8 |  |  |
| 2 | 15.4 | 22.2 |  | 14.8 | 21.1 |  |  |
| ≥3 | 67 | 46.2 |  | 74.5 | 55 |  |  |
| Zip-code wise median<br>income (in \$) (%) | | | 0.46 | | | 0.68 | <0.001 |
| 1 - 45,999 |  |  |  |  |  |  |  |
| 46,000 - 58,999 | 17.7 | 18.5 |  | 23.1 | 24.4 |  |  |
| 59,000 - 78,999 | 24 | 25.5 |  | 27 | 25.6 |  |  |
| 79,000+ | 28.4 | 27.3 |  | 26.7 | 26.7 |  |  |
|  | 29.7 | 28.5 |  | 23 | 23.2 |  |  |
| Co-morbidities (%) |  |  |  |  |  |  |  |
| COPD | 21.8 | 18.7 | 0.006 | 31.7 | 27.1 | 0.0006 | <0.001 |
| Stroke | 19.8 | 13.4 | <0.001 | 15.9 | 10.6 | <0.001 | 0.008 |
| Hypertension | 12.3 | 19.6 | <0.001 | 12.1 | 19.7 | <0.001 | 0.87 |
| PVD | 9.8 | 9.5 | 0.81 | 8.8 | 7.9 | 0.28 | 0.37 |
| Diabetes | 33.9 | 27.3 | <0.001 | 48.9 | 43.2 | 0.0003 | <0.001 |
| Obese | 11.1 | 10.9 | 0.81 | 27.4 | 27.7 | 0.81 | <0.001 |
| **Heart failure** | 78.6 | 70.9 | <0.001 | 79.5 | 71.9 | <0.001 | 0.55 |
| **CKD** | 49.6 | 31.5 | <0.001 | 50.6 | 30.1 | <0.001 | 0.59 |
| **Liver disease** | 2.6 | 1.3 | 0.0002 | 8.5 | 4.8 | <0.001 | <0.001 |
| **Hemodialysis** | 1.7 | 1 | 0.01 | 4.2 | 5.2 | 0.15 | <0.001 |
| **Smoking** | 32.9 | 35.4 | 0.09 | 32.4 | 38.1 | 0.0002 | 0.77 |
| **Anemia** | 48.2 | 27 | <0.001 | 49.3 | 29.1 | <0.001 | 0.55 |
| **Insurance (%)** |  |  | 0.42 |  |  | 0.008 | <0.001 |
| **Medicare** | 95.8 | 95.4 |  | 82.8 | 84.5 |  |  |
| **Medicaid** | 0.2 | 0.4 |  | 4.4 | 2.6 |  |  |
| **Private** | 3.6 | 3.9 |  | 11.9 | 12.1 |  |  |
| **Other** | 0.3 | 0.1 |  | 0.6 | 0.7 |  |  |
| **Hospital region (%)** |  |  | 0.76 |  |  | 0.98 | 0.004 |
| **Northeast** | 24.3 | 22.9 |  | 18.9 | 19.2 |  |  |
| **Midwest** | 22.3 | 22 |  | 22.1 | 22.5 |  |  |
| **South** | 32 | 32 |  | 38.4 | 37.8 |  |  |
| **West** | 21.2 | 22.1 |  | 20.4 | 20.3 |  |  |
| **Hospital Bed-size (%)** |  |  | 0.93 |  |  | 0.08 | 0.19 |
| **Small** | 6.6 | 6.9 |  | 6.4 | 8 |  |  |
| **Medium** | 20.9 | 20.9 |  | 18.1 | 19.8 |  |  |
| **Large** | 72.3 | 72 |  | 75.4 | 72.1 |  |  |
| **Location/teaching status of hospital (%)** |  |  | 0.88 |  |  | 0.06 | 0.07 |
| **Rural** | 0.7 | 0.8 |  | 0.2 | 0.7 |  |  |
| **Urban non-teaching** | 9 | 9.1 |  | 8.1 | 9 |  |  |
| **Urban teaching** | 90.2 | 90 |  | 91.6 | 90.2 |  |  |
| **Died (%)** | 5.7 | 0.3 | <0.001<sup>#</sup> | 4.3 | 0.2 | <0.001<sup>#</sup> | 0.01<sup>*</sup> |
| **Length of stay (in days)** | 7.2 | 2.6 | <0.001<sup>#</sup> | 8.7 | 2.8 | <0.001<sup>#</sup> | 0.03<sup>*</sup> |
| **Total charges (\$)** | 283,618 | 193,473 | <0.001<sup>#</sup> | 300,624 | 196,224 | <0.001<sup>#</sup> | 0.17<sup>*</sup> |

**Figure 3:**
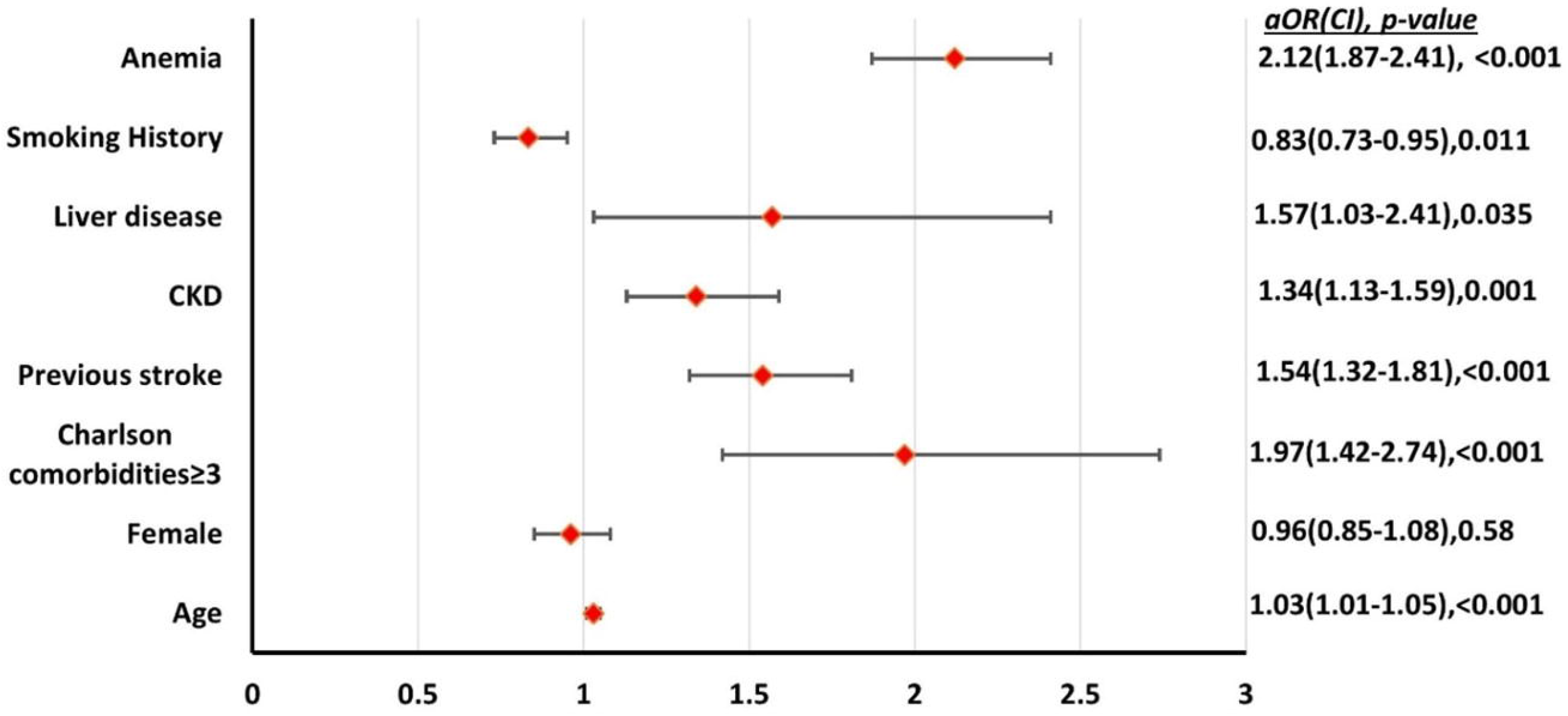
Independent factors associated with MAE in patients ≥80 years undergoing TAVR. CKD: Chronic Kidney disease

### 3.3. Comparison of TAVR patients <80 years with and without MAE

The mean age for patients with and without MAE was 71.2(SE±0.19) vs 71.4(SE±0.13) respectively (<0.001). The proportion of patients with comorbidities was significantly higher among those who experienced MAE compared to those without MAE (Table 3). Anemia (aOR-1.93), liver disease(aOR-1.48), CKD (aOR-1.68), history of stroke(aOR-1.46), multiple comorbidities were independently associated with higher odds of MAE. African American (aOR-0.69) race was associated with lower odds of MAE (Figure 4).

**Figure 4:**
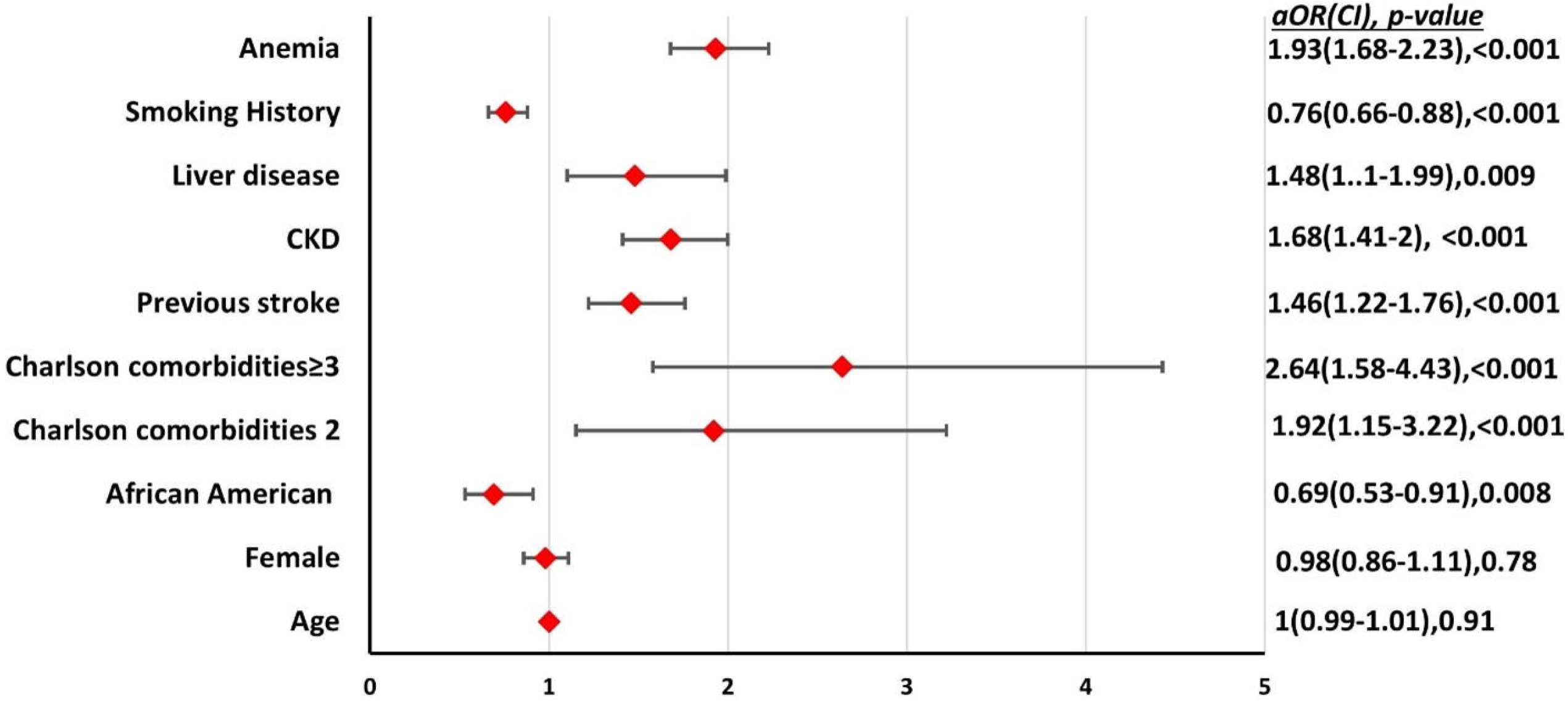
Independent factors associated with MAE in patients <80 years undergoing TAVR. CKD: Chronic Kidney disease.

### 3.4. Comparison of TAVR patients with MAE and age ≥80 years vs < 80 years

Among patients with in-hospital MAE, those < 80 years had higher comorbidities compared to those ≥80 years (Charlson category ≥3 - 74.5 vs 67%, p<0.001). More patients of age ≥80 years old also belonged to zip-codes with higher median income (p<0.001). On multivariate analysis, patients ≥80 years had higher in-hospital mortality compared to those less than 80 (5.7 vs 4.3%, aOR=1.58 [CI=1.08-2.32], p=0.01). The length of stay was also shorter for those ≥80 years (7.2 vs 8.7 days, p=0.03). There was no significant difference in the total hospital charges ($283,618 vs $300,624, p=0.17) between the two groups.

## 4. Discussion

Our study shows that patients undergoing those ≥80 years old had a higher in-hospital mortality rate compared to those < 80 years. There was no significant difference in the rates of MAE between the two cohorts. Anemia, CKD, Liver disease, and previous stroke were associated with higher odds of MAE in both groups.

Irrespective of the indication for admission, studies have shown increased in-hospital complication rates associated with older age resulting in worse outcomes (12). TAVR has been shown to be a relatively safe procedure even in nonagenarians (13). Our study shows that age over 80 years is associated with 1.5 times higher odds of in-hospital mortality. These results are of particular interest, given previous studies on similar comparisons have shown conflicting data. In their study of 1386 TAVR patients, Buellesfeld et al showed no difference in mortality among four age groups ranging from 40-99 years (14). Havakuk et al in their study comparing patients older than and less than 85 years reported no significant difference in the in-hospital and 30-day mortality between the two groups but noticed higher mortality in the older group on follow-up (15). Yamamoto et al in their comparison of patients >90 to <90 showed a trend of higher mortality in the older groups albeit not statistically significant (16). In their study of the TVT registry, Arsalan et al showed higher in-hospital death was among nonagenarians (6.5% vs. 4.5%; p <0.001)(17). Considering that patients ≥ 80 years had a lower proportion of comorbidities in our study compared to their younger cohorts, further risk stratification models may be needed to assess the factors influencing mortality in older patients undergoing TAVR.

Our study identified that anemia was associated with higher odds of in-hospital MAE in all sub-groups. TAVR patients with anemia ≥80 years old had marginally higher odds of MAE compared to those <80(a0R-2.12 vs 1.93). Anemia has been reported to be present in 30% of the patients with AS (18). Several studies have shown that anemia has been associated with worse short and long-term outcomes in patients undergoing TAVR(19-24). A meta-analysis by Kanjanahattakij et al showed increased long-term outcomes but no change in short-term mortality (25).

Our data also showed that liver disease was associated with higher odds of MAE in both ≥80 years and < 80 years old. The older subgroup had lower prevalence but slightly higher odds (a0R-1.48 vs 1.57) of MAE with liver disease. previous studies have shown that liver disease is associated with higher mortality and morbidity in patients undergoing TAVR. A multicenter study by Tirado-Conte et al showed that long-term non-cardiac mortality was higher in those with Liver disease, especially those with Child B and C cirrhosis, but in-hospital mortality (7%) was not affected by liver disease(26). In their retrospective study of 640 patients, Wendt et al showed that patients with Liver cirrhosis undergoing TAVR had an in-hospital mortality rate of 36.4%(27). The possible variation could be due to the difference in the severity of liver disease which cannot be assessed from the NIS database.

Our data shows that TAVR patients < 80 years old have a higher comorbidity burden compared to age ≥80 years, except for stroke and PVD. The possible explanation for this could be the wide use of risk stratification tools for patients undergoing TAVR, namely the STS (Society of Thoracic Surgeons) and euro SCORE II models. These models place older patients with AS and multiple comorbidities at unacceptably higher risk of adverse outcomes, thereby resulting in reduced TAVR rates in this population (28, 29).

Pacemaker insertion is a common procedure-related complication of TAVR (30). Despite the rapid technological advances in TAVR procedure, conduction abnormalities continue to be a significant complication requiring pacemaker insertion. Our study shows patients ≥80 had higher post-TAVR pacemaker insertion rates compared to those <80 years of age. Although not evaluated in this study, pre-existing conduction abnormalities and type of device have been known to be the strongest predictors of requiring a pacemaker. Increasing age has also been independently shown to be predictive of the need for PPM after TAVR(30). As a result of cardiac remodeling, older age is well known to be associated with conduction abnormalities, including bradyarrhythmia and tachyarrhythmia even in the absence of underlying heart disease. This is likely the causative factor for higher PPM insertion rates in this population (31).

Our study has several limitations. Firstly, data from the NIS is subject to biases associated with retrospective studies. Given that data is interpreted from NIS based on ICD codes, errors with coding may affect data accuracy. Additionally, due to the inherent design of NIS, long-term follow-up of outcomes is not possible. Since laboratory and pharmacological data are unavailable, utilization and comparison with STS and EUROSCORE were not feasible. Furthermore, comparing the rates of pacemaker insertion cannot be done since most studies report 30-day rates of pacemaker insertion rather than in-hospital rates. Finally, the NIS does not include information about the severity of the diagnosis at the time of admission. For example, the NYHA stage of heart failure or stage of CKD cannot be assessed.

## Conclusion

Overall, our study shows that in patients undergoing TAVR, the in-hospital mortality was slightly higher in those ≥80 years old compared to patients < 80 years. However, the rates of MAE were not significantly different between the two groups. Further prospective studies are required to build risk stratification models for older patients who undergo TAVR

## Data Availability

All data produced in the present study are available upon reasonable request to the authors

## Abbreviations

NIS: National inpatient sample
ICD-10-CM: using the International Classification of Diseases, Tenth Revision, Clinical Modification
TAVR: Transcatheter aortic valve replacement
MAE: major adverse events
LOS: length of hospital stay
THC: total hospital charges

