## Supplemental Table 1 for "Comparison of in-hospital mortality and clinical outcomes among patients greater than and less than 80 years undergoing TAVR"

### Table S1. TAVR procedure code.

|  | ICD-10 procedure code |
| --- | --- |
| Transcatheter Aortic Valve Replacement (TAVR) | 02RF37Z, 02RF38Z. 02RF3JZ, 02RF3KZ, 02RF37H, 02RF38H, 02RF3JH, 02RF3KH |

### Table S2. 10-CM codes for in-hospital major adverse events.

| **Adverse Events** | **ICD-10-CM codes** |
| --- | --- |
| **Post procedural hemorrhage** | D7821, D7822, D7831, D7832, E89810, E89811, E89820, E89821, G9751, G9752, G9761, G9762, H59311, H59312, H59313, H59319, H59321, H59322, H59323, H59329, H59331, H59332, H59333, H59339, H59341, H59342, H59343, H59349, H9541, H9542, H9551, H9552, I97610, I97611, I97618, I9762, I97620, I97621, I97630, I97631, I97638, J95830, J95831, J95860, J95861, K91840, K91841, K91870, K91871, L7621, L7631, M96830, M96840, N99820, N99821, N99840, N9984 |
| **Cardiac complications** | I21, I22, I46, I97710, I97790, I9788, I9789, I312, I314, I442, 0W9D30Z, 0W9D3ZX, 0W9D3ZZ, 0W9D40Z, 0W9D4ZX, 0W9D4ZZ, 02HK3J, 02HK3MZ, 02HL3JZ, 02HL3MZ, 02H63JZ, 02H73JZ, 02HK3JZ, 02HL3JZ, 5A1213Z, 5A1223Z, 0JH60PZ, 0JH60PZ, 0JH63PZ, 0JH63PZ, 0JH80PZ, 0JH80PZ, 0JH83PZ, 0JH83PZ, 0JH604Z, 0JH634Z, 0JH804Z, 0JH834Z, 0JH605Z, 0JH635Z, 0JH805Z, 0JH835Z, 0JH606Z, 0JH636Z, 0JH806Z, 0JH836Z, T8111XA,0 28D0ZZ, 02QD0ZZ, 02890ZZ, 02Q90ZZ, 02QG0ZZ, 02QH0ZZ, 02QJ0ZZ, 02BK0ZZ, 02NK0ZZ, 02NL0ZZ, 02QF0ZZ, 02QA0ZZ, 02B50ZZ, 02RM0JZ, 02U50JZ, 02UM0JZ, 02U50JZ, 02RM0JZ, 02UM0JZ, 02QF0ZZ, 02QG0ZZ, 02QH0ZZ, 02QJ0ZZ, 02U50JZ, 02UM0JZ, 02RM07Z, 02RM0KZ, 02U507Z, 02U508Z, 02U50KZ, 02UM07Z, 02UM0KZ, 02U507Z, 02U508Z, 02U50KZ, 02RM07Z, 02RM0KZ, 02UM07Z, 02RK07Z,0 2RK0KZ, 02RL07Z, 02RL0KZ, 02U607Z, 02U608Z, 02U707Z, 02U708Z, 02U70KZ, 02UK0KZ, 02UL0KZ, 02Q50ZZ, 02QM0ZZ, 02Q50ZZ, 02QM0ZZ, 02QB0ZZ, 02QC0ZZ, 02U50JZ, 021609P, 021609Q, 021609R, 02160AP, 02160AQ, 02160AR, 02160JP, 02160JQ, 02160JR, 02160KP, 02160KQ, 02160KR, 02160ZP, 02160ZQ, 02160ZR, 02W50JZ, 02WF07Z, 02WF08Z, 02WF0JZ, 02WF0KZ, 02WG07Z, 02WG08Z, 02WG0JZ, 02WG0KZ, 02WH07Z, 02WH08Z, 02WH0JZ, 02WH0KZ, 02WJ07Z, 02WJ08Z, 02WJ0JZ, 02WJ0KZ, 02WM0JZ, 02Q50ZZ, 02QM0ZZ, 021K0Z5, 021L0Z5, 02B60ZZ, 02B70ZZ, 02BK0ZZ, 02BL0ZZ, 02B60ZZ, 02B70ZZ, 02BK0ZZ, 02BL0ZZ |
| **Post-procedural stroke or transient ischemic attack** | G45, H340, H341, H342, I63 |
| **Acute kidney injury** | N170, N171, N172, N178, N179, N990, R34 |

ICD-10-CM: International Classification of Diseases, Tenth Revision, Clinical Modification.

### Table S3. Charlson Comorbidity Index and ICD-10-CM codes.

| **Comorbidity** | **Points** | **ICD 10-CM codes** |
| --- | --- | --- |
| Myocardial infarction | 1 | I25.2 |
| Congestive heart failure | 1 | I09.81, I25.5, I42.0, I42.5 - I42.9, I43.x, I50.x |
| Peripheral vascular disease | 1 | I70.x, I71.x, I72.x, I73.1, I73.8, I73.9, I77.1, I77.7, I79.0, I79.1, I79.8, I79.2, K55.1,  K55.8, K55.9, Z95.8, Z95.9 |
| Cerebrovascular disease | 1 | I69.x, Z86.73 |
| Dementia | 1 | F01.5, F02.8, F03.9, G30.x, G31.1, G31.8, G31.9 |
| Chronic pulmonary disease | 1 | I27.8, I27.9, J40.x - J47.x, J60.x - J67.x, J68.4, J70.1, J70.3, J84, J96.1 |
| Rheumatic disease | 1 | L94.0, L94.1, L94.3, M05.x, M06.x, M08.x, M12.0, M12.3, M30.x, M31.0 - M31.3,  M32.x - M35.x, M45.x, M46.5, M46.1, M46.8, M46.9, M48.8, M49.8 |
| Peptic ulcer disease | 1 | K25.5, K25.7, K25.9, K26.5, K26.7, K26.9, K27.5, K27.7, K27.9, K28.5, K28.7, K28.9 |
| Mild liver disease | 1 | B18.x, K70.0 - K70.3, K70.9, K71.3 - K71.5, K71.7, K73.x, K74.x, K76.0, K76.0, K76.4,  K76.8, K76.9, Z94.4 |
| Diabetes without chronic  complication | 1 | E08.9, E09.9, E10.9, E11.9, E13.9 |
| Diabetes with chronic complication | 2 | E08.2-E08.8, E09.x, E10.2 - E10.8, E11.2 - E11.8, E12.2 - E12.8, E13.2 - E13.8 |
| Hemiplegia | 2 | G04.1, G11.4, G80.1, G80.2, G81.x, G82.x, G83.0 - G83.5, G83.8, G83.9, I69.x, R53.2 |
| Renal disease | 2 | I12.0, I13.1, N18.x, N19.x, N25.0, Z49.0 - Z49.2 |
| Cancer | 2 | C00.x - C26.x, C30.x - C34.x, C37.x - C41.x, C43.x, C45.x - C58.x, C60.x - C76.x, C81.x  - C85.x, C88.x, C90.x - C96.x |
| Moderate or severe liver disease | 3 | I85.0, I86.4, K72.1, K72.9, K76.5, K76.6, K76.7 |
| Metastatic solid tumor | 6 | C77.x - C80.x, R18.0 |
| AIDS | 6 | B20 |

ICD-10-CM: International Classification of Diseases, Tenth Revision, Clinical Modification. AIDS: acquired immune deficiency syndrome.
